# High rate triggers increased atrial release of BMP10, a biomarker for atrial fibrillation and stroke, which affects ventricular cardiomyocytes

**DOI:** 10.1101/2025.02.24.25322820

**Authors:** LC Sommerfeld, J Schrapers, KF Müller, L Bravo, B Siebels, AMS Vermeer-Stoter, B Pan, G Höppner, C O’Shea, J Ridder, H Wieboldt, P Sander, T Zeller, W Chua, Y Purmah, RS Gardner, NR Tucker, P Kirchhof, MN Hirt, T Eschenhagen, J Stenzig, L Fabritz

**Affiliations:** University Center of Cardiovascular Science UCCS, University Medical Center Hamburg-Eppendorf, Martinistr. 52, 20246 Hamburg, Germany; University Heart and Vascular Center Hamburg, Department of Cardiology, Martinistr. 52, 20246 Hamburg, Germany; DZHK (German Center for Cardiovascular Research), partner site Hamburg/Kiel/Lübeck, Martinistr. 52, 20246 Hamburg, Germany; Institute of Experimental Pharmacology and Toxicology, University Medical Center Hamburg-Eppendorf, Martinistr. 52, 20246 Hamburg, Germany; Cancer and Genomic Sciences, University of Birmingham, B15 2TT Birmingham, United Kingdom; Cardiovascular Sciences, University of Birmingham, B15 2TT Birmingham, United Kingdom; Center of Diagnostics, Section Mass Spectrometry and Proteomics / Core Facility Mass Spectrometric Proteomics, University Medical Center Hamburg-Eppendorf, Martinistr. 52, 20246 Hamburg, Germany; Sandwell and West Birmingham Hospitals NHS Trust, B66 2QT Smethwick, UK; SUNY Upstate Medical University, Department of Pharmacology, 766 Irving Avenue Syracuse, NY 13210

## Abstract

**Background:** Bone morphogenetic protein 10 (BMP10) is a ligand of the TGFβ superfamily secreted mainly by atrial cardiomyocytes. Elevated BMP10 blood concentrations predict atrial fibrillation (AF), AF recurrence after ablation and AF-related cardiovascular complications like stroke. The conditions increasing BMP10 secretion and downstream effects of BMP10 in cardiomyocytes are poorly understood. We assessed BMP10 secretion dynamics and BMP10 effects in a human 3D model of atrial and ventricular engineered heart tissue (EHT).

**Methods:** Cardiomyocytes differentiated from human induced pluripotent stem cells (atrial and ventricular) were cast into a fibrin-matrix to generate EHT. Atrial EHTs were optogenetically paced (3-5 Hz) or maintained at intrinsic beating rate for 24 h up to 15 days. Release of BMP10 and other cardiac biomarkers from EHT were quantified. BMP10 plasma concentrations were compared between 1370 patients in different atrial rhythm at blood draw. Additionally, ventricular EHTs were exposed to BMP10 for 10 days.

**Results:** Atrial but not ventricular EHT released BMP10 within 48 h of culture. High rate optogenetic pacing increased atrial EHT BMP10 release by ∼3-fold after a latency of at least 24 h post pacing initiation. BMP10 plasma concentrations were elevated in patients with documented AF compared to sinus rhythm and even higher in patients with current AF. BMP10 induced upregulation of TGFβ pathway transcripts, increased expression of genes related to AF and heart failure, including *PITX2 and NPPB*, and increased relative contraction times in ventricular EHTs.

**Conclusions:** High atrial rates increase BMP10 expression and release, and BMP10 blood concentrations are higher in patients with current AF than in AF patients in sinus rhythm. High BMP10 concentrations induce expression of AF- and heart failure-related transcript networks in ventricular EHT. These findings support a role of BMP10 as a biomarker for AF and identify BMP10 as a potential player in AF-induced remodeling and tachycardiomyopathy.

**Clinical perspective:** *What is known?:*

- Bone morphogenetic protein 10 (BMP10) is a secreted member of the TGFβ-superfamily that is expressed in the heart.
- Elevated blood cof BMP10 are associated with AF, recurrent AF, and with AF-related complications such as stroke.

*What the study adds:*

- High atrial rates lead to BMP10 release from engineered atrial cardiac tissue.
- BMP10 activates cardiac expression of genes associated with AF and heart failure including *PITX2* and *NPPB*.
- BMP10 can play a role as a contributor to atrial cardiomyopathy and potentially to AF-induced ventricular dysfunction, specifically tachyarrhythmia-induced cardiomyopathy.

Graphical abstract:
High rate triggers increased atrial release of BMP10, a biomarker for atrial fibrillation and stroke, which affects ventricular cardiomyocytes.
BMP10: bone morphogenetic protein 10; EHT: engineered heart tissue. Created in https://BioRender.com.

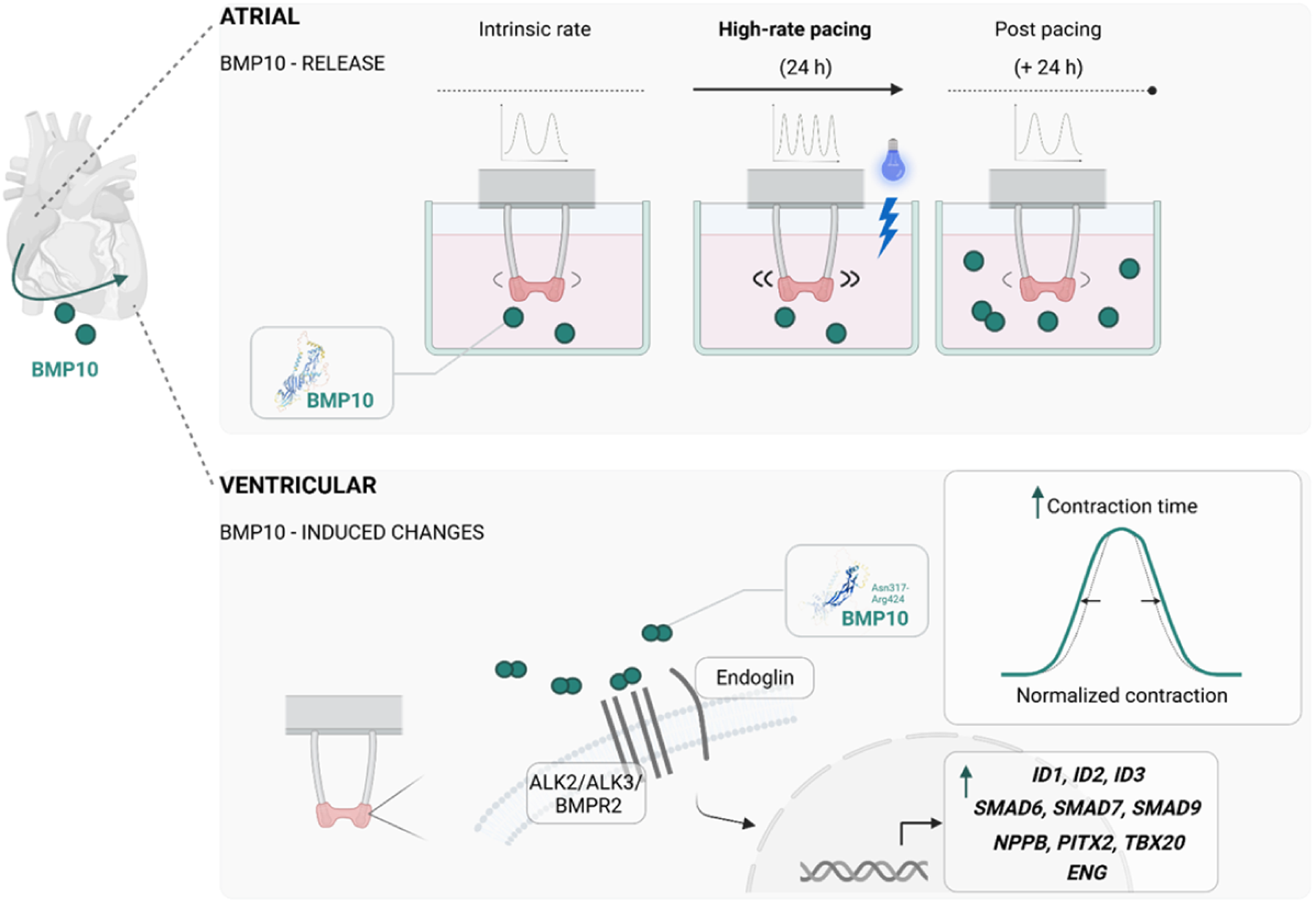

## Introduction

Atrial fibrillation (AF) affects 60 million individuals worldwide.^1^ Despite improved means of detection, many patients remain undiagnosed.^2^ AF itself leads to remodeling of the atria, thereby promoting its progression and puts patients at increased risk of stroke or heart failure. Early initiation of rhythm-control therapy can prevent AF-related complications including cardiovascular death, stroke, and heart failure hospitalizations^3^ and is recommended in current European and American guidelines.^4,5^ Quantifiable markers of AF burden would simplify effective early rhythm control.

Recently, bone morphogenetic protein 10 (BMP10) was identified as a putative blood biomarker for AF. Elevated blood concentrations of BMP10 were shown to be associated with AF^6^, stroke in patients with AF^7^, with recurrent AF after ablation^8,9^ and lower chance of attaining sinus rhythm at follow up.^10^ BMP10 in combination with other biomarkers can detect patients with prevalent AF but in sinus rhythm at medical contact^6^ and can identify patients with AF at high risk of cardiovascular events.^11^ Understanding the conditions leading to BMP10 release from the atria and the functional effects of BMP10 in the heart could help to better define the clinical use of BMP10.

BMP10 is a cardiac-specific secreted protein detectable in blood. BMP10 is expressed during mammalian embryogenesis and affects cardiomyocyte development and growth.^12^ In the adult heart *BMP10* expression is restricted to the atria.^8^

Despite its implication as an AF-associated biomarker, triggers for increased BMP10 expression and subsequent release are not known. Cardiac signaling resulting from BMP10 exposure in adulthood and its possible contribution to AF pathogenesis have not been studied. We investigated BMP10 release and downstream signaling in human cardiomyocytes (CM) using atrial and ventricular engineered heart tissue (aEHT/vEHT) as human *in vitro* models.^13,14^

## Methods

### EHT generation, culture and contractility analysis (aEHT/vEHT)

EHT were generated using previously established protocols.^13,15^ Both atrial and standard, ventricular cardiomyocytes (aCM/vCM) were differentiated from established human induced pluripotent stem cell lines of healthy donors. All cell lines employed in this work have been published (UKEi001-A, UKEi003-C), detailed information is available on https://hpscreg.eu. Cardiac differentiation was carried out following published protocols for both, aCM^13^ and vCM^15^. Fibrin-based EHT in 24-well cell culture format was created from 1×10^6^ aCM/vCM per EHT. After the onset of spontaneous beating activity at approximately 5-10 days after casting, contractility (force, frequency, rhythmicity, contraction and relaxation velocity) was analyzed every 2-3 days, 1 h after culture media change utilizing pattern recognition software (EHT Technologies and Consulting Team Machine Vision, CTMV, Pforzheim, Germany) following established protocols.^13,15^

Short-term electrical pacing for contractility analysis was carried out with custom-manufactured carbon electrodes (EHT Technologies).^14,16^ EHTs were paced at 1.25-2.25 Hz with biphasic impulses (4 ms, 2.25-4 V/cm, Grass S88X stimulator, Natus Neurology Incorporated). Pacing stimulus frequency and electrical field strength were titrated for each batch and set just above pacing threshold and spontaneous beating frequency. Averaged contraction peaks across treatment conditions were visualized for comparison by peak alignment of normalized traces, and the mean contraction as a function of time calculated.

### Optogenetic fast pacing of aEHT

To enable longer-term optogenetic pacing (3-5 Hz), the light-sensitive non-selective cation channel CheRiff2.0 was employed. Atrial EHTs were transduced during casting with an adeno-associated virus vector (AAV6) conferring channel expression under the control of a cardiomyocyte-specific cTNT promoter. A custom-manufactured LED-bearing circuit board (designed and manufactured by Julius Hansen, Sarcura GmbH, Klosterneuburg, Austria; circuit print by JLCPCB, Hong Kong, China) was placed underneath the 24-well cell culture plate harboring the EHTs. LEDs were addressed to rhythmically elicit blue light impulses (465 nm, 0.12 mW/mm^2^, 45 ms stimuli) by an Arduino controller (Nano ATMega328) using Arduino IDE software.

### Protein concentration analysis in EHT media

BMP10 in culture media was quantified by ELISA according to manufacturer’s instructions (EK18108, Human BMP-10 ELISA kit PicoKine, Boster, Pleasanton, USA). Fresh EHT culture media was confirmed to not contain any detectable BMP10. High sensitive troponin I (hsTnI), N-terminal pro-brain natriuretic peptide (NT-proBNP) and glucose media concentrations were measured using Abbott Architect Assays (Abbott Diagnostics) on Architect i2000 and c8000 Immuno Assay Analyzers (Abbott Diagnostics).

### BMP10 concentration analysis in patient blood

Blood BMP10 concentrations in patients were determined from the Birmingham and Black Country Atrial Fibrillation Registry (BBC-AF), which recruited consecutive patients from the Sandwell and West Birmingham NHS Trust (UK) between September 2014 and February 2018, as reported.^6,17^ Patients either had confirmed AF or other cardiovascular conditions (without AF) based on the CHA₂DS₂-VASc score; those without prior AF underwent 7-day ECG monitoring. The study was ethically approved (IRAS ID 97753), and complied with the Declaration of Helsinki. All participants provided written informed consent. Blood samples were immediately processed, stored at −80 °C, and analyzed centrally as reported before, using a high-throughput, high-precision assay (Roche Diagnostics, Penzberg, Germany).^6,17^ Clinical characteristics of patients by rhythm are listed in Supplementary Table S1.

### BMP10 exposure of vEHT

Effects of BMP10 on ventricular EHT (vEHT) contractility were assessed by both acute (30 min) and longer-term (10 days) exposure of vEHT to recombinant human BMP10 (rhBMP10, 2926-BP-025, R&D Systems, Minneapolis, USA) dissolved in water containing 0.1% bovine serum albumin and 4 mM HCl (vehicle).

For acute experiments, vEHTs were exposed to accumulating concentrations of rhBMP10 for 30 min each. Contractility was analyzed as described above. To allow for detection of potential positive inotropic effects, in contrast to standard EHT culture employing 1.8 mM extracellular Ca^2+^, extracellular Ca^2+^ was reduced to 1 mM.

### Gene expression analysis

RNA was extracted from either 31-38 day-old vEHT, or 35 day-old aEHT, with TRIzol reagent (Invitrogen, Carlsbad, USA). For RT-qPCR, reverse transcription was carried out with the High-Capacity cDNA Reverse Transcription kit (Applied Biosystems, Thermo Fisher, Vilnius, Lithuania) and qPCR using HOT FIREPol EvaGreen qPCR Mix Plus (Solis Biodyne, Tartu, Estonia). Primer sequences can be found in Supplementary Table S2.

RIN >9.2 was confirmed for all samples on an Agilent Tapestation device. Library preparation was carried out with the NEBNext Ultra II Directional RNA kit (New England Biolabs) and libraries were sequenced in paired-end mode (Nextseq 2000, Illumina), read length 111 bp, ∼25 million reads per sample.

A detailed description of cell type deconvolution of the bulk RNA sequencing data can be found in the supplement.

### Mass spectrometry-based proteomic analysis of EHTs

Please refer to the supplement.

### Statistical methods

For statistical analysis not related to RNA sequencing or mass spectrometry, GraphPad Prism software (v.6.0 and v.10.0, Dotmatics, Boston, USA) was used. Statistical tests are indicated in figure legends and were applied as appropriate: one-way or two-way ANOVA followed by Tukey’s or Šidák‘s multiple comparisons test. Sample numbers refer to n/n (number of EHTs/batches) and graphs depict individual values and mean±SEM unless stated otherwise.

## Results

### BMP10 expression and secretion is restricted to atrial EHT and increased with high rate pacing

To explore whether EHT is a suitable model to decipher BMP10 signaling mechanisms, we characterized both, aEHT and vEHT. *BMP10* mRNA was detectable in both models, although at ∼200-fold higher abundance in aEHT compared to vEHT (Figure 1A).

**Figure 1:**
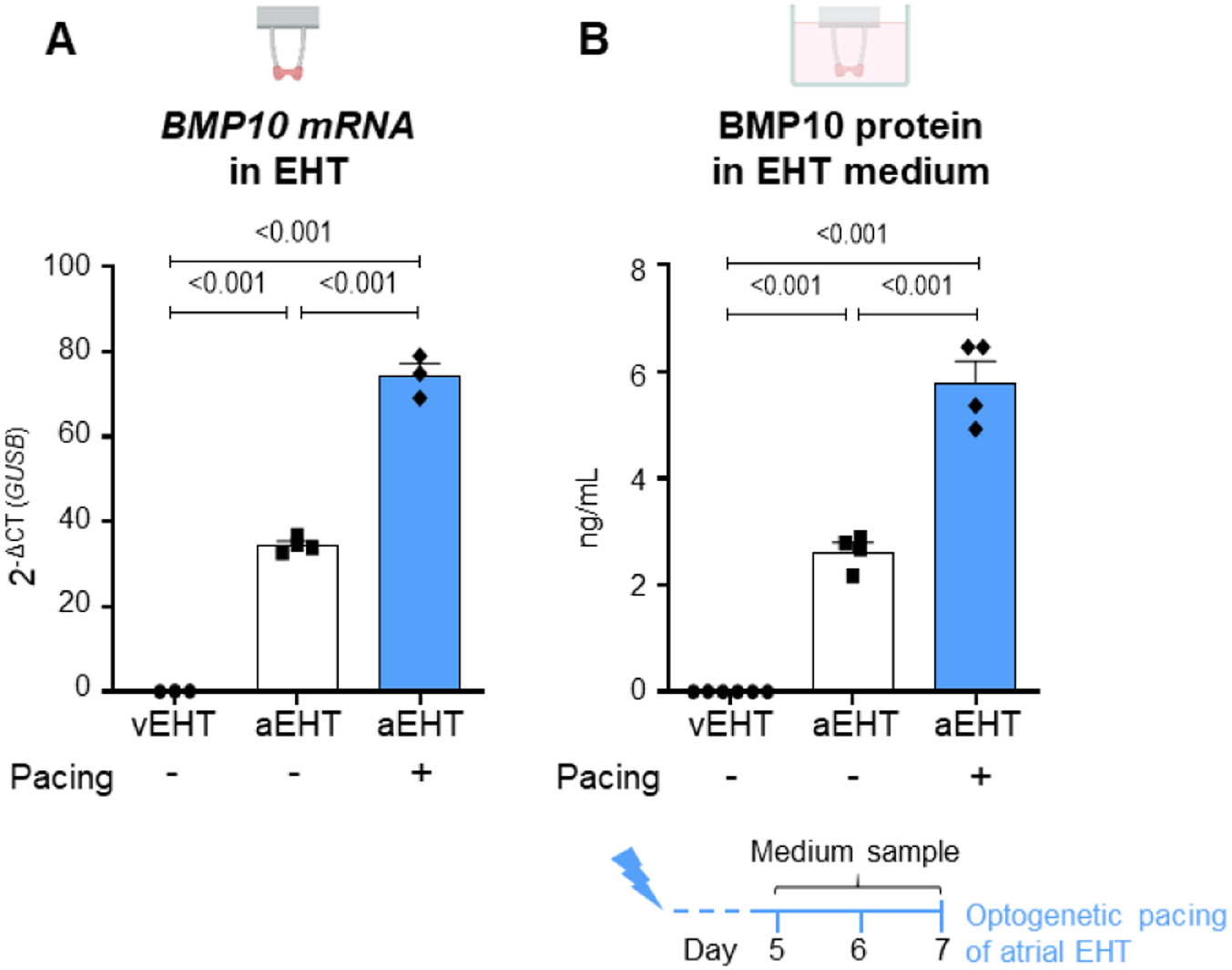
BMP10 expression and release by EHT. **(A)** *BMP10* mRNA abundance from spontaneously beating vEHT, spontaneously beating aEHT, and aEHT paced at 3 Hz for 7 days (+, blue, filled bars; RT-qPCR, n=3-4/1). **(B)** BMP10 in media from aEHT as in (A), collected after 48 h of aEHT culture (day 5-7, ELISA, n=4-6/1). One-way ANOVA followed by Tukey’s multiple comparisons test, p_adj_ reported for all comparisons.

Tachypacing of aEHT has been established as a model to study AF.^18,19^ To study the effect of high atrial rates on *BMP10* expression, we assessed *BMP10* mRNA concentrations in aEHTs optically continuously paced for 7 days at a frequency of 3 Hz. For control conditions, aEHTs were left to beat spontaneously at intrinsic beating frequency (usually 2 to 2.5 Hz). Pacing at 3 Hz increased *BMP10* mRNA expression in aEHT by 2-fold (Figure 1A). Average BMP10 concentrations in culture medium of aEHT were ∼2.6 ng/mL after 48 h in culture. In contrast, BMP10 was hardly detectable in vEHT media (Figure 1B). In line with higher gene expression, aEHT pacing led to a two-fold increase in BMP10 release (∼5.8 ng/mL within 48 h) compared to unpaced controls (Figure 1B).

### BMP10 secretion increases with rapid pacing, but with a delay

We paced aEHTs for 24 h at 5 Hz, followed by a post-pacing period of 24 h each, enabling time-course analysis of BMP10 secretion (Figure 2A). Optogenetic fast pacing for 24 h did not elevate BMP10 medium concentrations immediately. The increase in BMP10 by up to 1.7-fold was delayed to at least 24 h after pacing initiation (Figure 2B). By then, the protocol had reached the post-pacing period. Neither onset nor extent of BMP10 release showed rate-dependence within the studied pacing rate range of 3-5 Hz (Supplementary Figure S1). Pacing of aEHT led to an immediate increase in release of NT-proBNP and troponin I within 24 h (Figure 2C). Rapid pacing led to significantly increased glucose consumption (Figure 2C). Glucose consumption recovered to pre-pacing levels within 24 h. All observed effects were similar across 3, 4 and 5 Hz of applied rapid pacing (Supplementary Figure S1).

**Figure 2:**
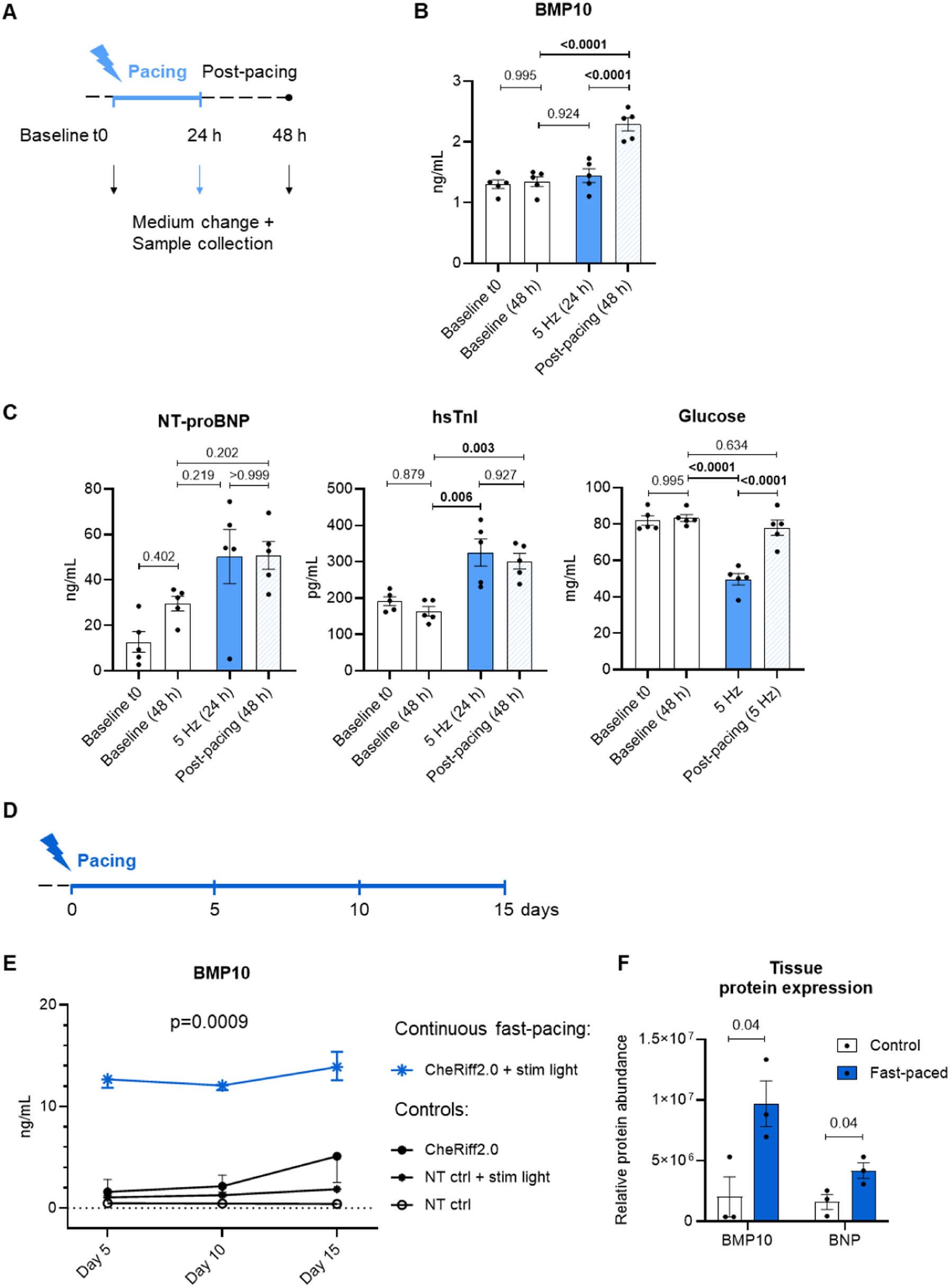
Media and tissue BMP10 content after continuous optogenetic fast pacing. **(A)** AEHTs were paced at 5 Hz for 24 h or left unpaced (B-C, n=5/1). First medium sample taken directly after pacing, second sample 24 h after the end of pacing (post-pacing) or 24 h after unpaced control culture. Quantification of **(B)** BMP10 and **(C)** high-sensitivity troponin I (hsTnI) as a marker for cell damage, N-terminal pro-brain natriuretic peptide (NT-proBNP) as a marker for cardiomyocyte stress and glucose for verification of fast pacing by higher glucose consumption in fast-beating EHTs. Data show glucose concentrations remaining after 24 h of culture. One-way ANOVA followed by Šidák’s multiple comparisons test. Adjusted p-values are reported. **(D)** AEHTs were continuously optically paced for 5, 10 and 15 days at 4.5 Hz (E-F). **(E)** BMP10 concentration in aEHT medium (accumulated during 48 h) throughout the protocol. Paired T-test, fast-paced vs controls. **(F)** Relative BMP10 and BNP protein abundance in aEHTs after culture, analyzed by mass spectrometry-based proteomics after 15 days of continuous fast pacing. Student’s T-test, n=3 per group. Contractility data for quality control can be found in the supplement (Supplementary Fig. S2). BNP: Brain natriuretic peptide; Ctrl: Control; NT: Non-transduced; t0: time point 0 at start of protocol. 24 h: 24 h after pacing initiation, 48 h: 48 h after pacing initiation and 24 h after switching back to intrinsic rate.

Continuous chronic pacing for up to 15 days (Figure 2D) led to consistently elevated BMP10 release from aEHT. After 5 days of continuous fast pacing at 4.5 Hz, BMP10 release was increased by ∼6-fold, without further increase until day 15 (Figure 2E). Long-term fast pacing caused increased protein expression of both, BMP10 and BNP (Figure 2F).

### BMP10 plasma concentrations are higher in patients with current AF

Plasma BMP10 concentrations differed between patients, depending on their rhythm status. Patients without AF and therefore in true sinus rhythm both at blood draw and during the consecutive week had the lowest BMP10 plasma concentrations, followed by those with a history of AF or Holter ECG-diagnosed AF within 7 days after the blood draw, but in sinus rhythm at the time of blood draw. Highest BMP10 plasma concentrations were observed in patients in AF on the day of blood draw (Figure 3A). In contrast, NT-proBNP concentrations did not differ between sinus rhythm patients and AF patients in sinus rhythm at blood draw (Figure 3B).

**Figure 3:**
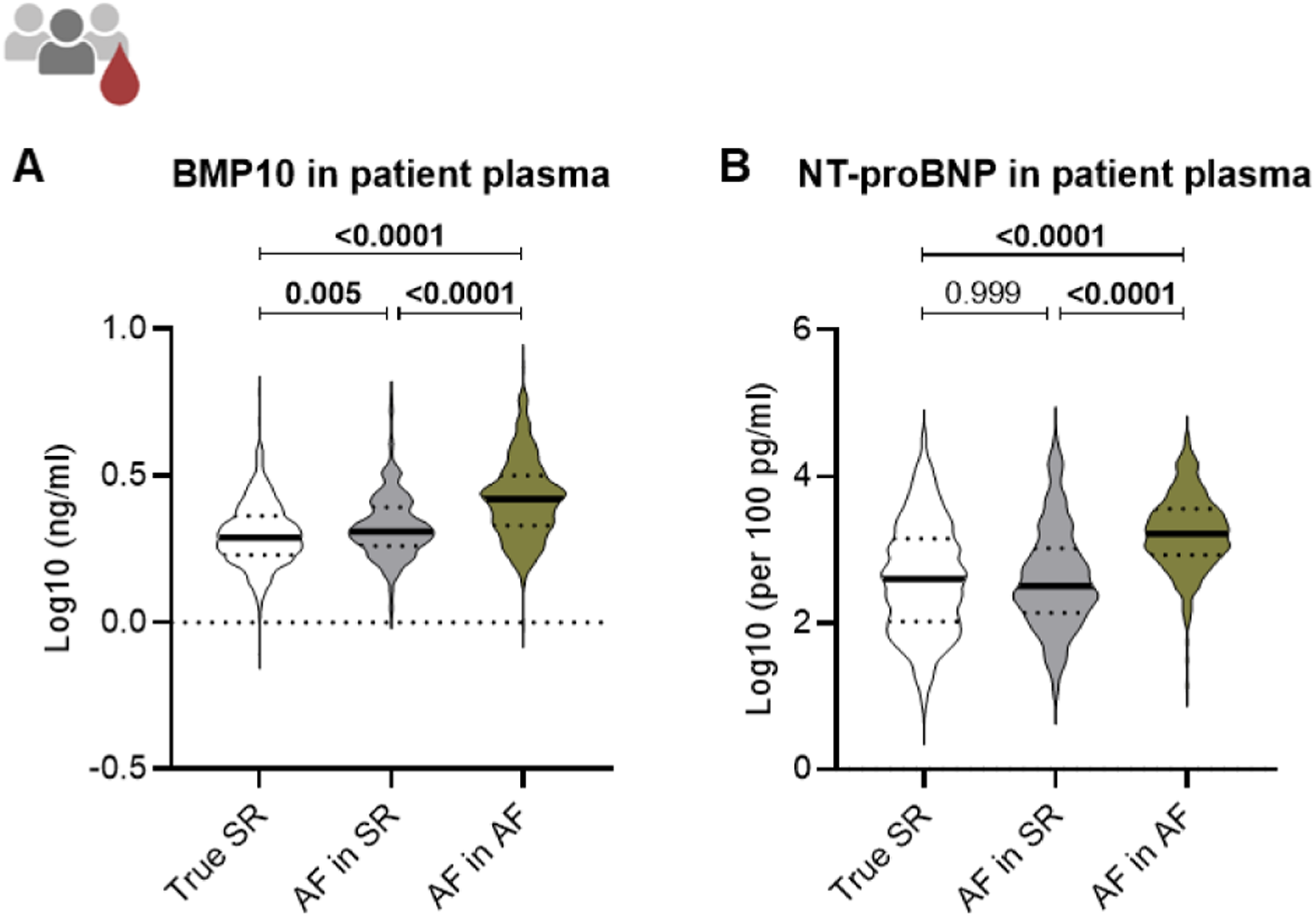
BMP10 and NT-proBNP plasma concentrations in patients by rhythm. Log10-transformed **(A)** BMP10 and **(B)** NT-proBNP plasma concentrations by rhythm at blood draw. Protein concentrations were quantified in plasma of patients without AF (True SR; sinus rhythm), with diagnosed atrial AF but in SR at the day of blood draw (AF in SR) and patients with AF in AF at the day of blood draw. n=True SR: 814, AF in SR: 254, AF in AF: 302. Median and quartiles are depicted. One-way ANOVA followed by Tukey’s multiple comparisons test, p_adj_ reported for all comparisons.

### Atrial EHT and vEHT express BMP10 receptors

Apart from endothelial cells and fibroblasts both, ventricular and atrial cardiomyocytes of the human adult heart express known BMP receptors (Figure 4A). To evaluate whether BMP10 can affect the human models employed here, we assessed the expression of BMP10 receptors in aEHT and vEHT. We identified *ALK3* and *BMPR2* as the known BMP receptors with highest expression in vEHT (Figure 4B) and confirmed expression of both receptors in aEHT (Figure 4C).

**Figure 4:**
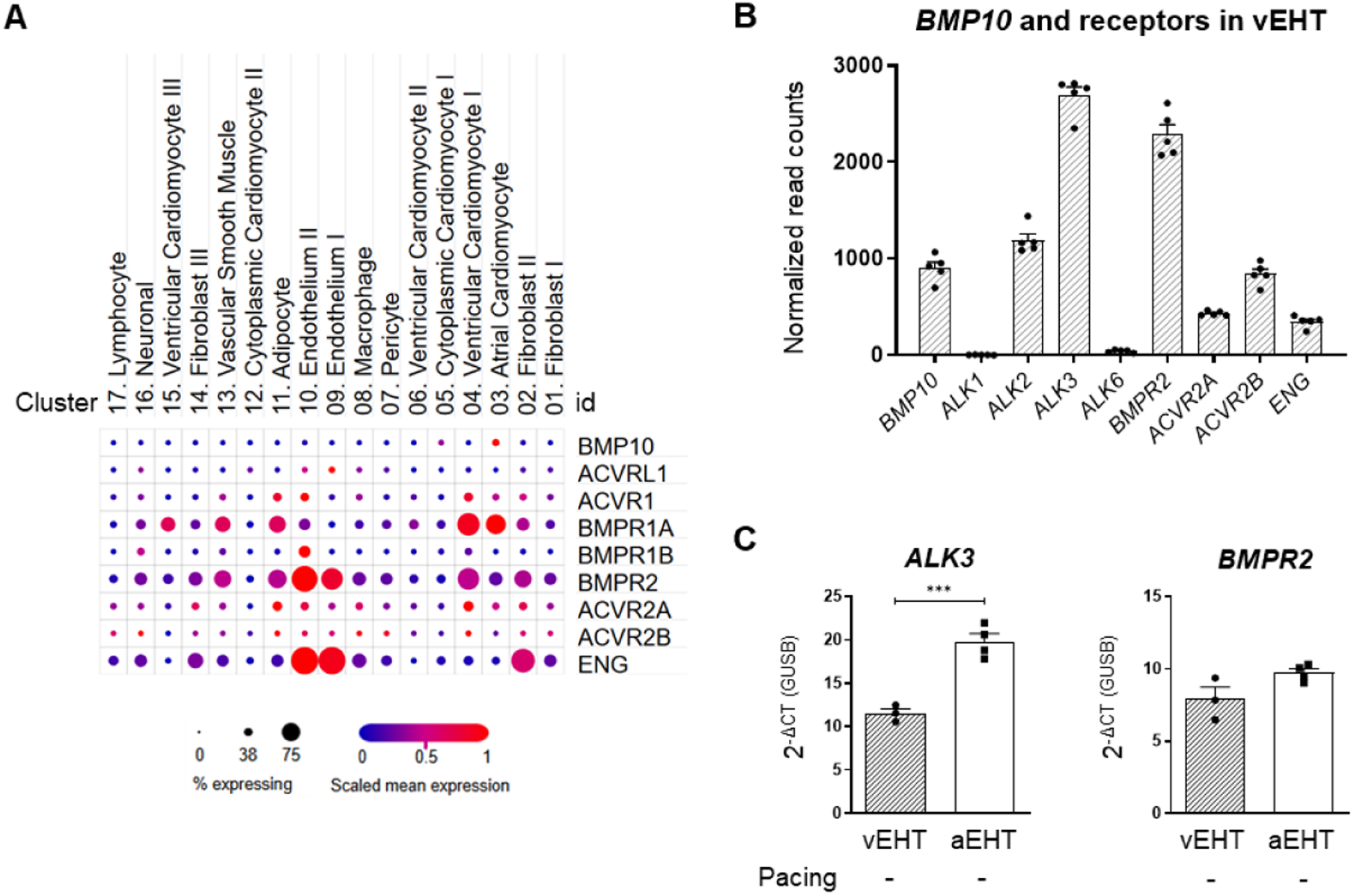
Expression of BMP10 and its receptors in vEHT and aEHT. **(A)** Expression of BMP10 and its known receptors in different cell populations of the adult human heart derived from snRNAseq analyses^20^, via https://singlecell.broadinstitute.org/single_cell. **(B)** BMP10 receptor expression in vEHT (RNA sequencing, n=5/1). **(C)** Relative mRNA abundance of *ALK3* and *BMPR2* in vEHT and aEHT, RT-qPCR, n=3-4/1. Unpaired T-test, ***p<0.001.

### BMP10 induces gene expression changes related to AF, driven by TGFβ signaling

The relevant abundance of BMP10 receptor transcripts in vEHT suggests that BMP10 could elicit effects on human myocardium and EHT. We therefore exposed vEHT to recombinant human BMP10 protein (rhBMP10) for 10 days. *BMP10* expression in vEHT was low at baseline, but we still observed a significant decrease in expression in reaction to rhBMP10 exposure. Other cardiac genes linked to atrial function and upregulated by rhBMP10 in a concentration-dependent fashion included *PITX2*, *NPPB* and *TBX20* (Figure 5A). Expression of the transcription factor *NKX2-5* was not affected by BMP10 exposure at the concentrations investigated.

**Figure 5:**
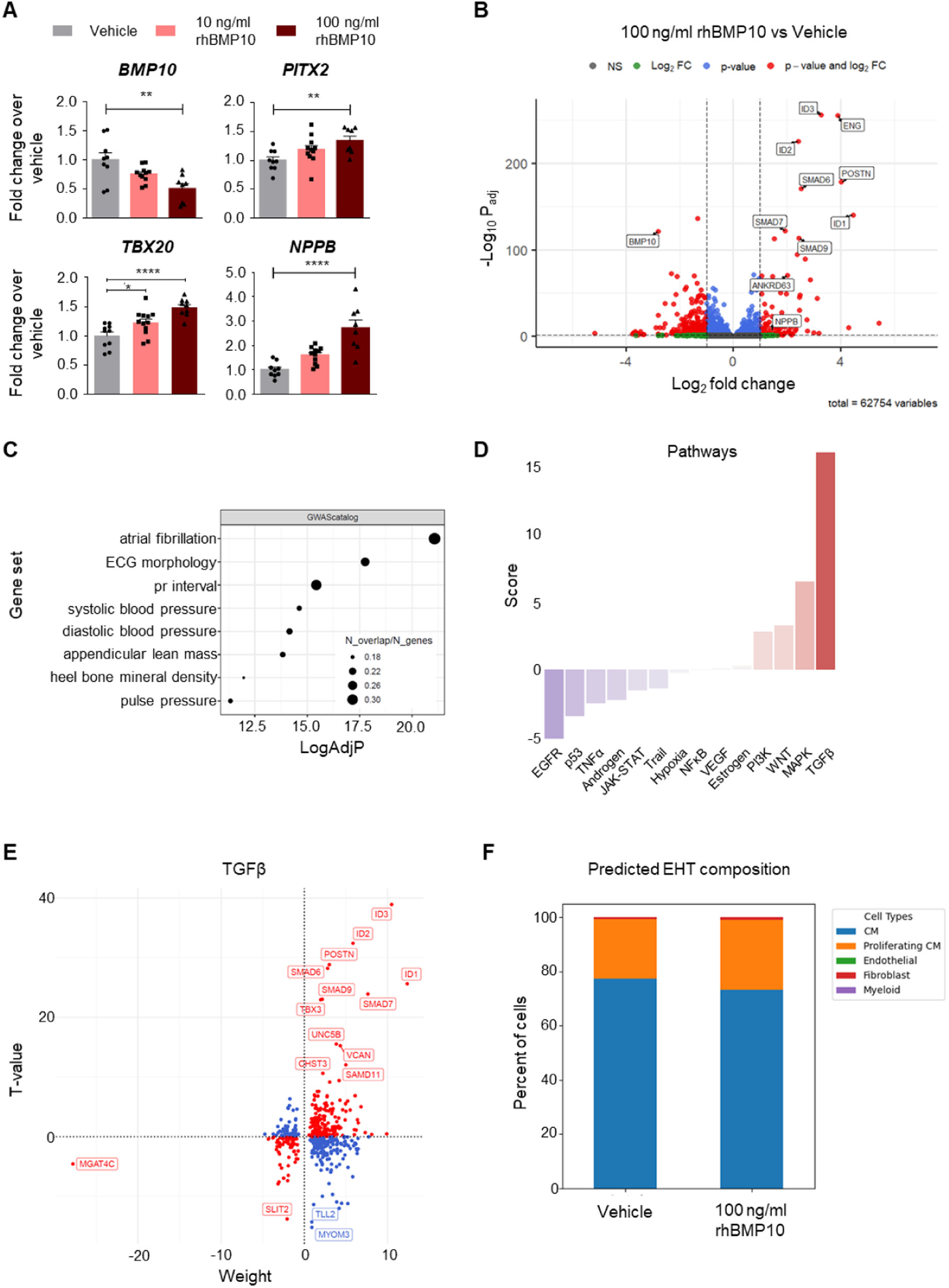
BMP10-induced gene expression shifts in ventricular EHT. **(A)** Gene expression of vEHT as assessed by RT-qPCR after 31-38 days of culture and 10 days of rhBMP10 exposure at 10 ng/mL and 100 ng/mL rhBMP10. 2^-ΔCT^ normalized to *GUSB* and vehicle group (n=8-12/2). One-way ANOVA followed by Bonferroni‘s multiple comparisons test, *p<0.05, **p<0.01, ****p<0.0001. **(B)** Bulk RNA sequencing results of 100 ng/ml rhBMP10-exposed vs. vehicle control vEHT (n=5/1) **(C)** Significantly enriched gene sets in differentially regulated transcripts. **(D)** Pathway analysis and **(E)** underlying expression changes in transcripts related to TGFβ signaling. Red indicates matching directionality of expression change in pathway-annotated transcripts. **(F)** Cell type content of vEHT as predicted by deconvolution analysis of bulk RNA sequencing data. Predicted EHT composition of individual EHTs can be found in supplemental material. CM: Cardiomyocyte; NS: Not significant; FC: Fold change.

These changes in expression of key atrial genes by chronic rhBMP10 exposure were confirmed in RNA sequencing analysis of vEHT exposed to 100 ng/mL rhBMP10 for 10 days (Supplementary Figure S3). Additionally, strongest expression increase induced by exposure to rhBMP10 was observed for ID genes (*ID1, ID2, ID3*), SMADs (*SMAD6, SMAD9*) and endoglin (*ENG*), a BMP10 receptor-encoding gene (Figure 5B). GWAS catalog analysis of all regulated transcripts showed strongest overlap with gene sets related to “atrial fibrillation”, “electrocardiogram morphology” and “PR interval” (Figure 5C). Pathway analysis revealed the highest score for the “TGFβ” pathway, driven mainly by ID genes, SMADs and *POSTN* (periostin, Figure 5D-E). Deconvolution analysis indicated that exposure to rhBMP10 did not affect cell type composition of the EHT (Figure 5F, Supplementary Figure S4).

### BMP10 exposure affects contraction time of vEHT

Functional consequences of rhBMP10 exposure in vEHT contraction were assessed acutely and chronically. Exposing vEHT acutely to increasing concentrations of rhBMP10 (0.5 ng/mL, 2.0 ng/mL, 10 ng/mL, 25 ng/mL, 100 ng/mL and 250 ng/mL) or vehicle for ∼30 min each did not affect contractility (Supplementary Figure S5).

Longer-term exposure to rhBMP10 for 10 days (100 ng/mL) resulted in increased time to peak (TTP −10%, −50% and −80%, Figure 6B, Supplementary Figure S6) as well as increased relaxation time (RT 10%, 50% and 80%, Figure 6B, Supplementary Figure S6), without affecting basal force (Figure 6A) or contraction and relaxation velocity (Figure 6C). Averaged, normalized contraction peaks depict the before-described rhBMP10-induced widening of the peak compared to control (Figure 6D).

**Figure 6:**
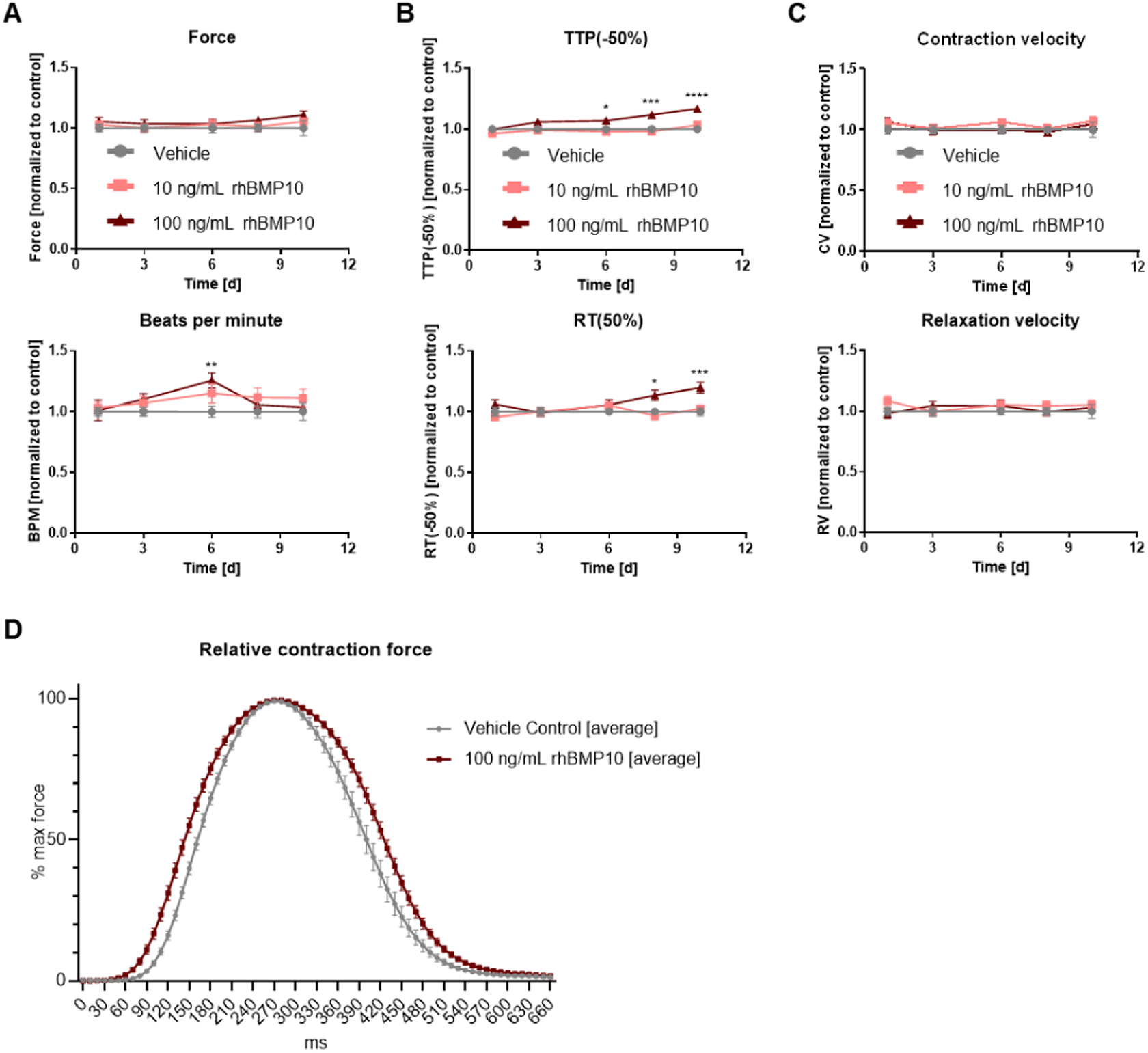
Contractility development of vEHT during longer-term exposure to rhBMP10. VEHTs were exposed to 10 ng/mL rhBMP10 or to 100 ng/mL rhBMP10 for 10 days (n=14-19/2). **(A)** Force development and beats per minute (BPM). **(B)** Time to peak (TTP; −50%) and relaxation time (RT; 50%) and **(C)** contraction (CV) and relaxation velocity (RV) are shown. TTP(−50%) refers to the time from 50% of maximal force to 100% maximal force. All values are normalized to vehicle group of respective batch. Two-way ANOVA followed by Šidák‘s multiple comparisons test, adjusted *p<0.05, **p<0.01, ***p<0.001, ****p<0.0001. **(D)** Averaged, normalized contraction peaks on day 10. Plotted are normalized mean contraction force as a function of time, aligned by peak.

## Discussion

This analysis of BMP10 release patterns in human atrial and ventricular engineered heart tissue (aEHT/vEHT) and BMP10 in patient plasma yielded the following findings (Graphical abstract):

I. BMP10 is expressed and secreted by aEHT.
II. High pacing rates increase BMP10 expression and release in aEHT.
III. Consistent with these findings, BMP10 plasma concentrations are most elevated in patients with current AF during blood draw. BMP10 concentrations are higher in patients in intermittent AF, but in sinus rhythm at blood draw, than in patients in true sinus rhythm.
IV. VEHT express BMP receptors and exposure to BMP10 leads to TGFβ-related gene expression with AF and heart failure.
V. BMP10 prolongs relative contraction and relaxation times in vEHT.

### Fast atrial rates increase BMP10 release from aEHT

This study shows that high atrial rates increase BMP10 secretion in engineered atrial tissue. Consistent with this finding, patients with AF at the time of blood sampling show higher BMP10 concentrations than patients with intermittent AF who are in sinus rhythm when blood is drawn (Figure 3). A decrease of BMP10 and NT-proBNP concentrations was seen after restoration of sinus rhythm.^21,22^

Unlike the increased glucose demand and the increased release of troponin and natriuretic peptides, BMP10 release occurred with a delay of at least 24 h. This delayed release supports a counter-regulatory function of BMP10, e.g. in maintaining cardiomyocyte metabolism^23^, proliferation^24^, or protection against cell death. The delayed but continuous BMP10 release after the tissue experienced high rates may correspond with increased BMP10 plasma concentrations in patients with intermittent paroxysmal AF but in sinus rhythm during the blood draw.

Optogenetic pacing of aEHT allowed us to study prolonged fast excitation in rates comparable to atrial rates in patients with AF without induction of pacing-related toxicity. Pacing increased metabolic demand, as evidenced by elevated glucose consumption, and triggered a stress response, reflected by increased BNP expression and release, as well as higher troponin concentration in the media. Notably, troponin and BMP10 were not released concurrently, arguing against the possibility that BMP10 increase in the media was merely a result of cell death and rupture. In patients with AF, both BMP10 and NT-proBNP (encoded by *NPPB*) plasma concentrations are elevated. In aEHT, pacing at high rates similarly led to increased BMP10 and NT-proBNP expression and secretion, aligning with the atrial biomarker profiles observed in AF patients.^10^ Ranging in the low ng/ml concentrations, the BMP10 concentrations detected in the aEHT media were comparable to those found in patient blood.

The pacing frequencies studied here (3-5 Hz) induce AF-related remodeling in aEHT within one week.^25^ It must be taken into account that aEHTs are chronically exposed to auto-secreted BMP10. We speculate that AF-related electrical remodeling observed in similar models is not solely attributable to tachypacing but may also result from the chronic autocrine and paracrine effects of BMP10 on the tissue.

The increase in BMP10 release appeared slower than that of NT-proBNP. This slow regulation, the atrial specificity and the stability over time in patients^22^ support the use of BMP10 as a marker for AF in patients. Since medium was changed every other day and levels remained stable, BMP10 was released continuously. This stability, coupled with its demonstrated ability to identify AF patients at higher stroke risk^7^, renders BMP10 a promising biomolecule for paroxysmal AF and AF-related complications. There is a clinical need to identify AF patient subpopulations at increased risk of complications, such as stroke. Recent clinical trials have revealed no correlation between anticoagulation efficacy and atrial high rate episodes (AHRE, now often referred to as “device-detected AF”) duration as a proxy for atrial arrhythmia severity.^26^ Current guidelines may leave some patient groups at risk of anticoagulation-related complications without deriving stroke prevention benefits. BMP10 offers potential as a biomarker for stratifying AF patients and addressing this unmet clinical need. Its detectability, even without ongoing AF, stability and association to endomysial fibrosis^27^ make it an attractive candidate for future research and clinical application.

EHT is a suitable and versatile model for studying cardiac biomarkers and the cardiac secretome in pathologies like AF, particularly BMP10 as reported here. Further studies on the temporal release pattern of BMP10 during AF initiation and progression are warranted.

### BMP10 impacts gene expression and contraction of ventricular EHT

BMP10, a growth factor of the TGFβ superfamily, is essential for ventricular trabeculation and maturation during cardiac development^12^ and maintains cardiomyocyte proliferation in adulthood.^24^ To evaluate whether BMP10 signaling can be activated in hiPSC-CMs, we examined BMP10 receptor expression in EHT. BMPs bind to different type I and type II serine/threonine kinase receptors and signal via the canonical SMAD-dependent nuclear pathway and several non-canonical pathways.^28^ BMP family proteins bind to the type I receptors activin receptor-like kinases (ALK) ALK1 (encoded by *ACVRL1*), ALK2 (*ACVR1*), ALK3 (*BMPR1A*) and ALK6 (*BMPR1B*)^29^ and form hetero-tetrameric signaling complexes with the type II receptors BMP receptor type II (BMPR2), activin receptor type 2A (ACVR2A) and activin receptor type 2B (ACVR2B).^30^ BMP receptor utilization is cell type-specific. Endoglin (ENG) acts as a co-receptor.^31^ *In vitro* analyses identified BMP9 and BMP10 to bind ALK1 with high affinity^32^ with ALK1 predominantly expressed in endothelial cells.^33^ In other cell types, BMP10 was observed to signal via ALK3 and ALK6.^34^ In the postnatal mouse heart, *Alk3* is expressed at much higher level than *Alk6*, suggesting ALK3 to be the main receptor for cardiac BMP10 signaling after development.^35^ In line with this, our results from vEHT indicate that BMP10 predominantly signals via ALK3 and BMPR2 in cardiomyocytes. Ventricular EHT provides a suitable model to explore effects of BMP10 exposure due to the low or undetectable intrinsic BMP10 expression, hence serving as an intrinsic control unaffected by varying endogenous BMP10 concentrations. This is important, as BMP10 levels will differ across cardiac regions. For instance, in a canine model, tachypacing at 600 bpm for seven days reduced BMP10 expression in the left atrium (LA, right atrium (RA) not studied).^36^

In vEHT, prolonged BMP10 exposure (10 days) upregulated *NPPB*, a key gene expressed in ventricular hypertrophy and heart failure. Elevated BMP10 concentrations have also been found in heart failure^37^, probably including patients with concomitant AF and HF.

BMP10 also increased expression of *TBX20*, a transcription factor essential for heart development and previously described as directly regulated by BMP10.^38^ Similarly, ID (Inhibitor of DNA binding) genes, known transcriptional regulators in early heart development^39^, were upregulated. Among these, ID2 is implicated in the specification of ventricular myocytes into conduction system lineages^40^ and associated with PR interval regulation in GWAS studies.^41^

BMP10 exposure did not alter the cell type composition of EHT. This indicates that the BMP10-induced effects are not attributable to differentiation into non-cardiomyocytes. However, due to BMP receptor expression in other cell types of the heart (Figure 3A), further studies assessing BMP10 effect on endothelial cells and fibroblasts in the heart are warranted.

Longer-term exposure of vEHT to rhBMP10 for 10 days led to changes in contractile properties, with effects becoming evident after six days. We observed a progressive delay in contraction and relaxation, resulting in a broadening of the contraction peak shoulders, without altering beating frequency or contraction force. Similar changes in gene expression and function have been reported during development of tachycardiomyopathy.^42,43^ These findings suggest that increased BMP10 secretion with high atrial rates contribute to arrhythmia-induced cardiomyopathy or, reflecting the prolonged contraction and relaxation times, to HFpEF, a common comorbidity in patients with AF. The effects of BMP10 compete with other effects such as oxidative and metabolic stress both in this model and in patients. Further research into the effects of BMP10 on cardiac function and its potential contribution to atrial cardiomyopathy^44^, to arrhythmia-induced cardiomyopathy and to the “vicious twins” of AF and heart failure is warranted.

## Data Availability

Data will be shared upon reasonable request to the corresponding author.

## Non-standard abbreviations and acronyms

BMP10: bone morphogenetic protein 10
EHT: engineered heart tissue

## Acknowledgments

We thank Dr. Ingke Braren, UKE Vector facility, for expert virus cloning and production. We thank Prof. Monika Stoll and Dr. Anika Witten, Core facility Münster, for RNA library preparation and sequencing.

## Author contributions

LF, JSt, LCS, and JSc designed the study. JSc and KFM performed EHT experiments and optogenetic pacing. HW, JR, LCS, and TZ conducted ELISA biomolecule measurements. LBM, LCS, RSG, and NRT performed RNAseq analysis. BS performed proteomic analysis. LCS, JSc, JSt, LBM, LF, WC, YP and CO’S analyzed the data. LCS, LBM, JSc, JSt, and LF interpreted the data. LF and JSt supervised the project. LF, PK, and LCS acquired funding. JSc, LCS, LF, and JSt wrote the manuscript. PK, TE and MNH critically reviewed the project. All authors have approved the final version of the manuscript.

## Sources of funding

Cofunded by UKE starter grant to LF and PK, by EU 633196 [CATCH ME] to LF and PK, EU 965286 [MAESTRIA] to LF, British Heart Foundation Accelerator Award (AA/18/2/34218) to University of Birmingham, Deutsches Zentrum für Herz-Kreislauf-Forschung (DZHK, German Center for Cardiovascular Research) supported by the German Ministry of Education and Research, including a postdoc start-up grant to LCS; German Research Foundation (Ki 509167694, STE2596/2-1, STE2596/4-1, STE2596/5-1, INST 337/15-1, INST 337/16-1, INST 152/837-1 and INST 152/947-1 FUGG).

## Disclosures

PK received research support from EZ, BHF, Leducq Foundation, MRC (UK), Else-Kröner-Fresenius-Stiftung, Dutch Heart Foundation and DZHK, from several drug and device companies active in atrial fibrillation and has received honoraria from several such companies in the past, but not in the last three years. PK is listed as inventor on two issued patents held by the employing institution (AF Therapy WO 2015140571, Markers for AF WO 2016012783). LF received institutional research grants by EU, BHF, DZHK, MRC (UK), NIHR and several biomedical companies active in the field of research. LF is listed as inventor on two issued patents held by the employing institution (AF Therapy WO 2015140571, Markers for AF WO 2016012783). BMP10 and NT-proBNP concentration in patient blood were provided by Roche as an in kind contribution as a partner in the CATCH-ME consortium.

## Supplemental Material

Supplementary Table S1-S2

Figures S1-S6

## Notes

### Competing Interest Statement

There is no direct competing interest, but we declare the following interests: LF received institutional research grants by EU 633196 [CATCH ME] and EU 965286 [MAESTRIA]. British Heart Foundation (AA/18/2/34218), German Center for Cardiovascular Research supported by the German Ministry of Education and Research (DZHK), Medical Research Council (UK), NIHR and several biomedical companies active in the field of research. LF is listed as inventor on two issued patents held by the employing institution (Atrial Fibrillation Therapy WO 2015140571, Markers for Atrial Fibrillation WO 2016012783). PK received research support for basic, translational, and clinical research projects from European Union, British Heart Foundation, Leducq Foundation, Medical Research Council (UK), Else-Kröner-Fresenius-Stiftung, Dutch Heart Foundation and German Center for Cardiovascular Research, from several drug and device companies active in atrial fibrillation and has received honoraria from several such companies in the past, but not in the last three years. PK is listed as inventor on two issued patents held by the employing institution (Atrial Fibrillation Therapy WO 2015140571

### Clinical Trial

Birmingham-Black Country-Atrial Fibrillation registry, IRAS ID: 97753

### Funding Statement

Cofunded by UKE starter grant to LF and PK, by EU [CATCH-ME] to LF and PK, EU 965286 [MAESTRIA] to LF, British Heart Foundation Accelarator Award (AA/18/2/34218) to University of Birmingham, Deutsches Zentrum für Herz-Kreislauf-Forschung (DZHK, German Center for Cardiovascular Research) supported by the German Ministry of Education and Research, including a postdoc start-up grant to LCS; German Research Foundation (Ki 509167694, STE2596/2-1, STE2596/4-1, STE2596/5-1, INST 337/15-1, INST 337/16-1, INST 152/837-1 and INST 152/947-1 FUGG).

### Author Declarations

Health Research Authority (HRA), United Kingdom

